# Comparison of Post-Extubation Pulmonary Outcomes Between High Flow Nasal Cannula and Conventional Oxygen Therapy in Elderly Cardiac Surgery Patients

**DOI:** 10.1101/2025.04.25.25326417

**Authors:** Suttasinee Petsakul, Jutarat Tanasansuttiporn, Wilasinee Jitpakdee, Pongsanae Duangpakdee, Thanapon Nilmoje, Panjai Choochuen, Wit Wichaidit, Chanikarn Boonyakiat, Sumidtra Prathep

## Abstract

**Background:** High-flow nasal cannula (HFNC) therapy is increasingly utilized following extubation in patients undergoing cardiac surgery, due to its potential benefits in preventing pulmonary complications such as atelectasis, pleural effusion, and pneumonia. However, the comparative efficacy of HFNC versus conventional oxygen therapy (COT) in elderly patients remains unclear. This study aims to compare post-extubation pulmonary outcomes between HFNC and COT in elderly patients following cardiac surgery.

**Methods:** We conducted a retrospective cohort study at a tertiary care hospital in Southern Thailand, including patients aged 65 and older who underwent elective cardiac surgery between December 2018 and January 2024. The primary outcome was the incidence of post-extubation pulmonary complications (PPCs) within 72 hours. Secondary outcomes included the length of intensive care unit (ICU) and hospital stays, all-cause in-hospital mortality, and atelectasis scores.

**Results:** A total of 424 patients were included, with 212 patients in each group. HFNC therapy was associated with a significant reduction in the odds of bronchospasm (7.5% vs. 24.5%; adjusted OR = 0.22, 95% CI = 0.12–0.41). There were no significant differences between the groups in terms of reintubation rates, tracheostomy, ICU stay, hospital stay, or all-cause mortality. However, atelectasis scores were significantly improved on day 5 post-extubation.

**Conclusion:** HFNC therapy may reduce the incidence of bronchospasms in elderly patients after cardiac surgery and significant improvement of atelectasis score at day 5. While these findings suggest a potential benefit of HFNC in postoperative care, further research is warranted to explore its long-term effects and implications for patient outcomes.

## INTRODUCTION

High-flow nasal cannula (HFNC) therapy has recently gained traction post-extubation intervention. One of its primary advantages over conventional oxygen therapy (COT) is the ability to generate continuous positive airway pressure (CPAP), which can facilitate partial lung recruitment^1^. This mechanism may reduce ventilation dead space, enhance oxygenation, and improve patient comfort in critically ill individuals^2^.

Postoperative pulmonary complications (PPCs) are significant risk factors for patients undergoing cardiac surgery^3^, contributing to increased morbidity and mortality, as well as prolonged stays in the Intensive care unit(ICU) and hospital. The incidence of pulmonary complications following cardiac surgery is notably high, affecting approximately 55% of patients^4^. Over the past decade, HFNC therapy has been increasingly used in the early postoperative phase to mitigate these complications. By generating low levels of positive airway pressure, HFNC can wash out upper airway dead space and aid in the clearance of secretions^5^. Additionally, characteristics of HFNC, such as increased FRC and end-expiratory lung volume (EELV), may improve oxygenation, especially in older patients^6^.

Despite these potential benefits, the evidence supporting HFNC use in elderly patients following cardiac surgery remains limited. A meta-analysis comparing HFNC with conventional oxygen therapy via face mask in adults extubated after cardiac surgery identified only two suitable studies, encompassing a total of 495 patients. While HFNC was associated with reduced “escalation of therapy”, the reintubation rates were similar between the two groups^7^. This meta-analysis highlights the limited evidence available and underscores the need for further research to investigate the benefits of HFNC after cardiac surgery, particularly in elderly patients who may derive unique advantages from this therapy. HFNC’s ability to maintain a constant FiO_2_ during peak inspiratory flow, provide low-level continuous positive airway pressure, increase end-expiratory volume, and reduce the work of breathing suggests it could be beneficial for geriatric patients.^8,9^

However, the effects of HFNC therapy following extubation in elderly patients at high risk of reintubation remain unclear. Previous studies, including one focused on elderly patients, have yielded inconclusive results regarding the benefits of HFNC^10^. Furthermore, the impact of HFNC in the context of cardiac surgery has not been thoroughly investigated. Given these gaps in literature, this study aims to compare post-extubation respiratory outcomes in elderly patients undergoing cardiac surgery at a tertiary facility in Southern Thailand. By addressing this understudied area, we seek to provide clearer evidence regarding the efficacy of HFNC in this vulnerable patient population.

## METHODS

### Study Design and Setting

This retrospective cohort study was conducted by analyzing data from electronic medical records at Songklanagarind Hospital, a tertiary care center in Southern Thailand. The study was designed to evaluate the effectiveness of different oxygen therapy methods in elderly patients following elective cardiac surgery.

### Study participants

The study population included patients aged 65 years and older who underwent elective cardiac surgery between 1 December 2018 and 31 January 2024. Patients was consecutive case collection. Patients were excluded if they had an ASA classification higher than Level 4, required preoperative ventilator support, or had a history of previous cardiac surgery. These criteria were selected to focus on relatively homogeneous population at significant risk of post-extubation pulmonary complications (PPCs).

### Sample Size Calculation

The sample size was calculated based on the anticipated incidence of reintubation within 72 h post-extubation. Assuming a reintubation rate of 0% in the HFNC group and 5% in the COT group^11^. We used R software with the epical package to estimate the required sample size. To detect a significant difference with 95% confidence level and 80% power, we determined that data from at least 190 patients per group were needed. Accounting for a potential 10% rate of missing data, we increased the sample size to 212 patients per group, resulting in a total of 424 patients.

### Oxygen Therapy Methods

We compared outcomes between patients receiving high-flow nasal cannula (HFNC) therapy and those receiving conventional oxygen therapy (COT). HFNC was delivered via a nasal cannula (Optiflow; Fisher & Paykel Healthcare Ltd, Auckland, New Zealand) connected to a humidifier with an integrated flow generator (AIRVO 2; Fisher & Paykel Healthcare Ltd, Auckland, New Zealand), providing warmed and humidified respiratory gases. COT was administered using a simple face mask or an oxygen mask, depending on the patient’s FiO2 requirements. The initial flow rate for both therapies was determined by the attending staff and adjusted as needed to maintain SpO2 between 92% and 98%.

### Outcomes: Clinical Outcomes of the Study Participants

The primary outcome was the incidence of PPCs within 72 hours post-surgery, including pneumonia, pleural effusion requiring thoracocentesis, pneumothorax, bronchospasm necessitating bronchodilators, and hypoxia (PaO_2_/FiO_2_ ≤ 100). Secondary outcomes included the duration of ICU stay, total length of hospital stay, all-cause in-hospital mortality, and changes in lung status as observed on chest radiographs on days 2 and 5 compared with baseline.

### Data Collection and Management

Following ethical approval, we accessed patient data through the hospital’s electronic medical record system. Eligible patients were identified and their data extracted using a standardized case record form (CRF). The CRF comprised two sections: (A) baseline characteristics, intraoperative and postoperative clinical characteristics, and (B) postoperative arterial blood gas values at multiple time points, including pre-extubation and post-extubation. The CRF also included an item for scoring atelectasis. Data were anonymized, with all personally identifiable information (patients’ names and hospital numbers (HN)) removed and entered into KoboToolbox for analysis.

### Statistical Analysis

All analyses were conducted using R version 4.2.2. Descriptive statistics were used to summarize baseline characteristics and outcomes. Continuous variables were compared between the HFNC and COT groups using parametric or nonparametric tests, depending on the distribution of the data. Categorical outcomes were compared using chi-squared or Fisher’s exact tests. To examine the association between oxygen therapy method and clinical outcomes, we performed bivariate, univariate, and multivariate logistic regression analyses. In the multivariate analysis, we adjusted for confounding variables, including age, sex, body mass index, smoking history, and chronic lung diseases, based on their known impact on respiratory outcomes^12,13^. All statistical analyses were performed at a 95% confidence level.

### Ethical Considerations

This study was conducted in accordance with the Declaration of Helsinki and was approved by the Human Research Ethics Committee of the Faculty of Medicine at the Prince of Songkla University, Songkhla, Thailand (Ethics Committee reference number: REC.66-526-8-1). Informed consent was not required because the data were retrospective and anonymized.

## RESULTS

Data were collected from 424 elderly patients, with 212 patients in each group (HFNC and COT). The mean age of participants was approximately 70 years in both groups (*Table 1*). The majority of participants were male and had comorbidities such as hypertension renal disease, liver cirrhosis, cardiovascular accident (CVA), diabetes, chronic lung disease, smoking history, with a relatively low prevalence of chronic lung disease. A notable difference between the groups was a higher proportion of current smokers in the HFNC group compared to the COT group. The Left ventricular ejection fraction (LVEF) was divided to good (>50%), moderate (31-50%), poor (21-30%) and very poor (<20%). There was no significant between HFNC and COT group. New York Heart Association (NYHA) and Euroscore were not significant between HFNC and COT group. The type of surgery, aortic cross-clamp time (AOX) and CPB time were not significant between HFNC an COT group. The duration of mechanical ventilator was shorter in HFNC group compared to COT group: 720 and 852.5 minutes respectively (p<0.001).

**Table 1.** Baseline characteristics.

|  | **COT (n=212)** | **HFNC (n=212)** | **P-value** |
| --- | --- | --- | --- |
| Age (year) | 70 (67, 75) | 71 (68, 76) | 0.114 |
| Male | 123 (58.0%) | 140 (66.0%) | 0.109 |
| BMI (kg/sqm) | 24.07 (21.93, 27.00) | 23.82 (21.70, 26.14) | 0.340 |
| **Comorbidities** |  |  |  |
| Hypertension (mmHg) | 159 (75.0%) | 159 (75.0%) | 0.999 |
| Renal disease on hemodialysis | 1 (0.5%) | 8 (3.8%) | **0.043\*** |
| Liver cirrhosis | 3 (1.4%) | 0 (0.0%) | 0.247 |
| CVA | 21 (9.9%) | 21 (9.9%) | 0.999 |
| Others | 17 (8.0%) | 20 (9.4%) | 0.731 |
| **DM** |  |  |  |
| Oral hypoglycemic agent use | 49 (23.1%) | 55 (25.9%) | 0.792 |
| Insulin use | 11 (5.2%) | 11 (5.2%) |  |
| **Chronic lung disease (Long term use bronchodilator or steroid use)** |  |  |  |
| COPD | 5 (2.4%) | 11 (5.2%) | 0.206 |
| Asthma | 9 (4.3%) | 6 (2.8%) | 0.592 |
| OSA | 2 (0.9%) | 5 (2.4%) | 0.450 |
| Any chronic lung disease | 15 (7.1%) | 20 (9.4%) | 0.489 |
| **Smoking** |  |  |  |
| Never | 121 (57.1%) | 95 (44.8%) | **0.021\*** |
| Former smokers (smoked in lifetime but not within 3 months of baseline) | 80 (37.7%) | 96 (45.3%) |  |
| Active (smoked within 3 months of baseline) | 11 (5.2%) | 21 (9.9%) |  |
| **LVEF (%)** |  |  |  |
| Good (>50%) | 139 (69.8%) | 135 (64.9%) | 0.299 |
| Moderate (31-50%) | 48 (24.1%) | 55 (26.4%) |  |
| Poor (21-30%) | 12 (6.0%) | 15 (7.2%) |  |
| Very poor (<20%) | 0 (0.0%) | 3 (1.4%) |  |
| **NYHA** |  |  |  |
| Class I | 18 (8.5%) | 25 (11.8%) | 0.548 |
| Class II | 106 (50.2%) | 98 (46.2%) |  |
| Class III | 85 (40.3%) | 85 (40.1%) |  |
| Class IV | 2 (0.9%) | 4 (1.9%) |  |
| **Euroscore** | 1.395 (0.975, 2.075) | 1.500 (0.970, 2.440) | 0.290 |
| **Type of surgery** |  |  |  |
| CABG | 114 (53.8%) | 122 (57.8%) | 0.459 |
| AVR | 42 (19.8%) | 32 (15.2%) | 0.259 |
| Mitral valve repair/replacement | 36 (17.0%) | 32 (15.2%) | 0.707 |
| CABG + Valve | 25 (11.8%) | 25 (11.8%) | 0.999 |
| Repair of structural defect | 0 (0.0%) | 1 (0.5%) | 0.998 |
| Resection of cardiac tumor | 1 (0.5%) | 1 (0.5%) | 0.999 |
| Others | 16 (7.5%) | 14 (6.6%) | 0.860 |
| AOX time (mins) | 63.50 (43.75, 94.25) | 68.00 (48.00, 103.25) | 0.114 |
| CPB time (mins) | 98.5 (74.75, 131.25) | 108.0 (79.5, 141.5) | 0.098 |
| Cryoprecipitate (ml) | 0 (0, 0) | 0 (0, 0) | 0.057 |
| Duration of mechanical ventilator (mins) | 852.5 (330, 1111.2) | 720 (220, 967.5) | **<0.001\*** |

(COT, conventional oxygen therapy; HFNC, high flow oxygen cannula; BMI, Body mass index; CVA, cardiovascular accident; COPD, chronic obstructive pulmonary disease; OSA, obstructive sleep apnea; LVEF, left ventricular ejection fraction; NYHA, New York Heart Association; CABG, Coronary artery bypass grafting; AVR, aortic valve replacement; AOX, aortic cross-clamp time; CPB, cardiopulmonary bypass) (values are expressed as mean ± SD or number (proportion) or median (IQR). \*P<0.05)

In terms of respiratory complications, we collected the data of pneumonia, pleural effusion, pneumothorax, bronchospasm and hypoxia. The incidence of bronchospasm was significantly lower in the HFNC group compared to the COT group (7.5% vs. 24.5%; adjusted OR 0.22; 95% CI = 0.12, 0.41) (*Table 2*). However, there were no significant differences between these two groups regarding secondary outcomes, including reintubation rates, tracheostomy rates, and all-cause mortality as shown in Table 3. Additionally, there were no significant differences in the length of hospital stay (10.1 ± 11.5 vs 10.2 ± 11.3 days) or ICU stay (4.4 ± 3.8 vs 4.1 ± 7.2 days) between the HFNC and COT groups (*Table 4*). Atelectasis scores demonstrated statistically significant improvements in the HFNC group compared to the COT group on days 5 compared to baseline (*Table 5*), indicating a more favorable respiratory recovery trajectory in patients receiving HFNC.

**Table 2.** Risk of pulmonary complications: COT vs. HFNC.

| **Complication** | **COT**<br>**(N= 212)**<br>**n (%)** | **HFNC**<br>**(N=212)**<br>**n (%)** | **Unadjusted OR**<br>**(95% CI)** | **Adjusted OR**<br>**(95% CI)** |
| --- | --- | --- | --- | --- |
| **Respiratory complication** |  |  |  |  |
| Pneumonia | 6(2.8) | 3(1.4) | 0.49 (0.12,2.00) | 0.29 (0.05,1.85) |
| Pleural effusion (needing<br>thoracentesis) | 4(1.9) | 7(3.3) | 1.78 (0.51,6.16) | 1.13 (0.24,5.31) |
| Pneumothorax | 4(1.9) | 5(2.4) | 1.26 (0.33,4.74) | 1.46 (0.36,5.93) |
| Bronchospasm (need<br>bronchodilator) | 52(24.5) | 16(7.5) | 0.25 (0.14,0.46) | **0.22 (0.12,0.41)** |
| Hypoxia | 22(10.4) | 16(7.6) | 0.71 (0.36,1.39) | 0.81 (0.38,1.71) |
| **Respiratory complication**<br>**(any type)** | 60(28.4) | 35(16.7)) | 0.50 (0.31,0.81) | **0.50 (0.30, 0.82)** |

2 Bold texts denote statistical significance at 95% level of confidence
\*Adjusted for age, sex, body mass index, smoking, and history of chronic lung.
4 (COT, conventional oxygen therapy; HFNC, high flow oxygen cannula)

**Table 3.** Comparison of secondary outcomes between COT and HFNC.

| **Intervention** | **COT**<br>**(N=212)**<br>**n (%)** | **HFNC**<br>**(N=212)**<br>**n (%)** | **Unadjusted OR**<br>**(95% CI)** | **Adjusted OR**<br>**(95% CI)** |
| --- | --- | --- | --- | --- |
| **Reintubation** | 8(3.8) | 10(4.7) | 1.27 (0.49,3.28) | 0.95 (0.32, 2.82) |
| **Tracheostomy** | 2(0.9) | 2(0.9) | 0.99 (0.14,7.13) | 0.61 (0.06,6.49) |
| **All-cause mortality** | 2(0.9) | 5(2.4) | 2.54 (0.49, 13.22) | 2.57 (0.43, 15.37) |

\*Adjusted for age, sex, body mass index, smoking, and history of chronic lung. (COT, conventional oxygen therapy; HFNC, high flow oxygen cannula)

**Table 4.** Comparison of length of hospital stays and ICU stay between COT and HFNC.

| **Outcome** | **COT**<br>**(N=212)**<br>**n (%)** | **HFNC**<br>**(N=212)**<br>**n (%)** | **Unadjusted Beta**<br>**(95% CI)** | **Adjusted Beta**<br>**(95% CI)** |
| --- | --- | --- | --- | --- |
| **Length of hospital stay** | 10.2 ± 11.3 | 10.1 ± 11.5 | -0.019 (-2.192, 2.154) | -0.228 (-2.436, 1.979) |
| **Length of ICU stay** | 4.1 ± 7.2 | 4.4 ± 3.8 | 0.358 (-0.736, 1.453) | 0.422 (-0.696, 1.540) |

(COT, conventional oxygen therapy; HFNC, high flow oxygen cannula)

**Table 5.** Atelectasis score among patients who received COT and HFNC.

| Characteristic | COT<br>(N=212)<br>n (%) | HFNC<br>(N=212)<br>n (%) | P-value* |
| --- | --- | --- | --- |
| Improved of atelectasis at<br>Day 2 vs. Baseline | 0(0) | 3(1.4) | 0.208 |
| Improved of atelectasis at<br>Day 5 vs. Baseline | 0(0) | 4(1.9) | <0.001 |

\* Fisher's test or chi-square test.
(COT, conventional oxygen therapy; HFNC, high flow oxygen cannula)

## DISCUSSION

In this hospital-based retrospective study, we compared respiratory and other clinical outcomes in elderly cardiac surgery patients who received either HFNC oxygen or COT. Our findings indicate that patients treated with HFNC had significantly lower odds of experiencing bronchospasm compared to those treated with COT. However, no significant differences were observed between the two groups concerning other respiratory outcomes and majority of secondary outcome. However, there was significantly improved of atelectasis score at day 5 compared to baseline. These results have important implications for practitioners in anesthesiology and elderly care.

Bypass surgery can lead to ischemic–reperfusion injury, impacting the intravascular compartment adjacent to the pulmonary microcirculation. Initially, “no reflow” was described in the coronary vasculature during embolism, where diminished blood flow and concurrent ischemia cause local endothelial and interstitial tissues to swell and protrude into the lumen, obstructing flow^14^. Postoperative wheezing is a potential complication in these patients. Moreover, secretion production decreases by age so elderly patient may be susceptible to develop mucus thickness with difficulty clearance which may resulting ineffective cough and bronchospasm^15,16^. Our study demonstrated that HFNC could reduce the incidence of bronchospasms and also improved of atelectasis score at day 5 postoperatively, a finding previously emphasized in studies focused on major abdominal surgery^17,18^. Research suggests that HFNC can create positive end-expiratory pressure (PEEP) by impeding expiratory flow, with pressure levels reaching 5–7.5 cm H_2_0. Non-invasive ventilation provides both oxygenation and positive end-expiratory pressure (PEEP), which can contribute to improved ventilation and potentially reduce the risk of bronchospasm and atelectasis. According to clinical practice guideline of intensive care physician recommended HFNC as moderate certainty on postoperative cardiothoracic surgery^19^. Our recent study was convincing the safety and efficacy of HFNC among these specific patient and operation. Furthermore, future studies should consider stratifying patients by specific disease patterns (i.e., obstructive vs. restrictive pulmonary disease) and clinical scenarios (e.g., hypercapnic respiratory failure, COPD, asthma exacerbations) to determine whether the observed benefits in respiratory mechanics translate into meaningful clinical improvements^20^. HFNC was report of barotrauma in specific condition mentioned above^21^ which may cause a difficulty for application HFNC as general patients.

Our study’s strengths include the completeness of the data and the absence of follow-up loss, which minimized the potential for selection bias. Although HFNC has become increasingly common, replacing COT as the standard method in recent years, the demographic characteristics and medical technologies influencing respiratory outcomes remained relatively stable during the study period. Therefore, we do not expect significant selection bias due to temporal factors. Additionally, since clinical procedures and outcomes were recorded by hospital staff according to standard practice guidelines, information bias is unlikely. We adjusted for all significant predictors of respiratory and secondary outcomes in our multivariate regression analysis, minimizing the risk of residual confounding.

Despite these strengths, certain limitations should be noted. First, our data were collected from a single tertiary care hospital in southern Thailand, which may limit the generalizability of our findings. The effects of HFNC and COT may differ in other health settings, particularly those with fewer intensive care staff. Second, we only measured outcomes up to 72 hours post-extubation, leaving long-term health outcomes unexplored. Future studies should expand the analysis to include different settings and longer-term follow-up to better understand the broader impact of these therapies.

## CONCLUSION

In conclusion, our study suggests that HFNC therapy may offer significant benefits in reducing post-extubation pulmonary complications, particularly bronchospasm and improve atelectasis score at day 5 post-operatively in elderly patients following cardiac surgery. These findings support the consideration of HFNC as a preferred option in postoperative care protocols, with an emphasis on careful monitoring and individualized patient management to optimize outcomes. Further research is warranted to confirm these results across diverse clinical settings, explore long-term outcomes, and refine the criteria for HFNC use in this patient population to maximize its clinical effectiveness.

## Data Availability

All relevant data are within the manuscript and its Supporting Information files.

## Notes

### Competing Interest Statement

The authors have declared no competing interest.

### Funding Statement

The author(s) received no specific funding for this work.

### Author Declarations

This study was conducted in accordance with the Declaration of Helsinki and was approved by the Human Research Ethics Committee of the Faculty of Medicine at the Prince of Songkla University, Songkhla, Thailand (Ethics Committee reference number: REC.66-526-8-1). Informed consent was obtained prior to data collection.

